# Differential fMRI neural synchrony associated with migraine during naturalistic stimuli with negative emotional valence

**DOI:** 10.1101/2024.12.20.24319445

**Authors:** Keva Klamer, Joshua Craig, Christina Haines, KiAnna Sullivan, Peter Seres, Chelsea Ekstrand

## Abstract

Migraine is a common neurological disorder that impacts approximately 12% of the general population and is characterized by moderate to severe headaches, nausea, mood changes, and fatigue. It impacts lower-level visual and auditory processing, causing hypersensitivities that lead to heightened audiovisual multisensory integration. However, the impact of migraine on the processing of complex, audiovisual stimuli is still unclear. Additionally, migraine may induce hypersensitivities to emotional arousal and valence, though the relative significance of these factors remains unknown. The current study seeks to identify how migraine impacts synchronous neural processing of complex, audiovisual stimuli, and how this differs based on the emotional arousal and valence of the stimulus. To do so, we collected functional magnetic resonance imaging data from 22 migraineurs and 21 healthy controls during the passive viewing of three audiovisual films of differing emotional arousal and valence. We identified that, in response to a negative valence, high arousal emotional stimulus, the migraine group showed greater neural synchrony in regions associated with multisensory integration, including the bilateral posterior superior temporal gyrus (pSTG), superior parietal lobule (SPL), and left middle temporal gyrus (MTG). There were no significant differences in neural synchrony between the migraine and control groups in response to positive valence, high arousal and neutral valence, low arousal stimuli. These findings suggest that migraine involves hypersensitivity to audiovisual movies as a function of negative emotional valence, where negative/aversive emotional states may drive greater synchrony in multisensory integration. Overall, this research highlights distinct pathways through which emotion and arousal impact neural processing in migraine.

---

Migraine is a common neurological disorder that impacts approximately 12% of the general population and is more common in females than males (Szabó et al., 2019). It is characterized by unilateral moderate to severe headaches, nausea, mood changes, and fatigue (Silberstein, 1995). Migraineurs experience enhanced perception and altered cerebral processing of sensory stimuli, commonly referred to as generalized hyperexcitable cortex. Hyperexcitable cortex refers to a reduction of intracortical inhibition, which leads to the excitability of neurons (Ciuffini et al., 2020) and is present even during the interictal period (i.e., in between migraine attacks). As a result, migraineurs experience hypersensitivities to lower-level stimuli (e.g., visual/auditory stimuli) and higher-level stimuli (e.g., painful/emotional stimuli; Wang et al., 2017). This suggests that migraineurs are producing a different neurological reaction than non-migraineurs when perceiving the same visual and auditory stimuli.

There are several different subtypes of migraine, as highlighted by The International Classification of Headache Disorders (3^rd^ edition; ICHD-3; Headache Classification Committee of the International Headache Society, 2013), which has been shown to display excellent specificity (Göbel et al., 2020). The ICHD criteria includes several subtypes of migraine, including migraine without aura, probable migraine without aura, migraine with aura, and probable migraine with aura. Migraine without aura is defined as a headache lasting 4-72 hours that may involve unilateral location, pulsating quality, moderate or severe pain intensity, aggravation by physical activity, and the headache may be accompanied by nausea and/or vomiting and photophobia and phonophobia. Probable migraine without aura is chararacterized by migraine-like attacks missing one of the features required to fill all criteria for a type or subtype of migraine, indicating that individuals with probable migraine without aura are on the spectrum of migraine disorders. Probable migraine without aura has symptom and epidemiologic profiles that overlap with migraine, and is therefore considered a prevalent form of migraine (Patel et al., 2004). Migraine with aura is defined as recurrent attacks of unilateral fully-reversible visual sensory or other symptoms that usually develop gradually and are followed by headache and associated migraine symptoms (Headache Classification Committee of the International Headache Society, 2013). Similarly, probable migraine with aura refers to aura-like episodes that are missing one criterion required for a full diagnosis of migraine with aura, but still share key clinical features (Headache Classification Committee of the International Headache Society, 2013). Individuals with probable migraine with aura are considered part of the broader spectrum of migraine with aura disorders. All of these conditions involve recurrent headache attacks with overlapping symptomatology and underlying pathophysiological mechanisms.

Migraine is associated with atypical unimodal processing of lower level stimuli during the interictal period, specifically auditory and visual hypersensitivities. In the domain of visual hypersensitivities, migraineurs have an increased sensitivity to normal light, flickering lights (e.g., computer screens), bright light (e.g., sunlight), and patterns (e.g., plaid, achromatic gratings; Friedman & De Ver Dye, 2009; Chong et al., 2016). Within the scope of auditory hypersensitivities, approximately 75% of migraineurs report sensitivities to sound during the interictal period (Vingen et al., 1998), and studies have shown that migraineurs are more sensitive to sound compared to healthy controls (Ashkenazi et al., 2009). Further, migraineurs exhibit lower light and auditory discomfort thresholds compared to controls (Meylakh & Henderson, 2022). These findings highlight that migraineurs experience atypical processing of both visual and auditory stimuli, leading to heightened sensitivity and lower discomfort thresholds during the interictal period.

Atypical unimodal processing of lower level stimuli during the interictal period in migraine has been increasingly linked to altered multisensory integration. Multisensory integration refers to the production of a single perception of the environment through the co-modulation of different modes of incoming stimuli (Schwedt, 2013). In other words, multisensory integration allows for the formation of a coherent perception of everyday life by combining inputs from different sensory modalities. Multisensory integration is a crucial component of daily lived experience, and several pieces of evidence suggest that migraineurs show altered multisensory integration compared to healthy controls (Schwedt, 2013). First, the presence of one migraine symptom (e.g., visual hypersensitivity) positively correlates with the intensity of other migraine symptoms (e.g., auditory hypersensitivity, pain severity; Kelman & Tanis, 2006). Second, exposure to visual, auditory, and olfactory stimuli can trigger migraine attacks (Harle et al., 2006). Third, exposure to one mode of sensory stimulation alters the sensitivity to concurrent sensory stimuli of other modalities (Ashkenazi et al., 2010). Fourth, studies have shown that audiovisual integration operates over a longer time window in migraineurs compared to controls (O’Hare, 2017) and that controls are better than migraineurs at motion discrimination, which involves integration of a stimulus over time (O’Hare, 2018). More specifically, research has shown that audiovisual integration is significantly elevated, and audiovisual suppression is weaker, in migraineurs compared to controls (Yang et al., 2014). This may be the result of visual and auditory hypersensitivities, which reflect an increased excitability state toward headache attacks (i.e., cortical hyperexcitability; Boran et al., 2021).

Several brain regions are implicated in the integration and coordination of visual and auditory inputs, and their involvement may vary depending on the complexity of the stimulus (Gao et al., 2023). These regions include the bilateral posterior superior temporal gyrus (pSTG), bilateral superior parietal lobule/intraparietal sulcus (SPL/IPS), right lingual gyrus, right thalamus, left middle temporal gyrus (MTG), and bilateral frontal gyri. The pSTG is considered the hub of multisensory integration, where it integrates auditory and visual speech (Ozker et al., 2017) and is activated during multisensory integration of simple and complex speech, and complex nonspeech stimuli (Gao et al., 2023). The SPL/IPS is more generally involved in multisensory integration as it processes both auditory and visual stimuli, whereby auditory inputs arrive before visual inputs (Molholm et al., 2006). In addition, the SPL shows greater activation in response to congruent stimuli across different senses (e.g., a visual cue that matches an auditory cue) compared to congruent stimuli within a single sense (e.g., two visual cues) during fMRI (Saito et al., 2005). The right frontal gyri are activated in response to simple stimuli, while the left MTG and frontal gyri respond to complex non-speech stimuli. In contrast, complex speech stimuli activate the right lingual gyrus, right thalamus, and left inferior occipital gyrus (Gao et al., 2023). These findings demonstrate the complex interplay of various brain regions in the integration of visual and auditory stimuli, where the pSTG consistently plays a central role, regardless of stimulus complexity.

Emotional features of stimuli can also influence multisensory processing. Emotional arousal, which refers to the intensity of an emotional response (Citron et al., 2014) has a relationship with multisensory processing, where multisensory stimulation has been shown to amplify the emotional arousal experienced when compared to unimodal stimulation (Schreuder et al., 2016). Further, high levels of emotional arousal may generally alter perception, where they are linked to increased attentional engagement (Zsidó, 2024). For example, research suggests that multisensory integration may be enhanced by positive emotional valence stimuli (Takeshima, 2020). In addition, brain regions involved in multisensory integration, such as the pSTG, show altered activation levels in response to emotionally charged stimuli, suggesting that emotional content can enhance or modify multisensory processing in these areas (Klasen et al., 2014). Further, neural network models designed for multisensory integration also show improved performance when emotional features are incorporated. However, negative emotional valence stimuli may slow or impair audiovisual multisensory integration (Taffou et al., 2017), suggesting a complex relationship between emotional valence, arousal, and multisensory integration.

In line with this, migraineurs show increased neural activation in the visual cortex and cerebellum to emotional static picture stimuli (e.g., emotional faces), and this heightened neural response becomes especially pronounced when the valence of the incoming stimulus is negative (Szabó et al., 2019; Wilcox et al., 2016). Therefore, it has been theorized that migraineurs may have a general hypersensitivity to aversive/negative stimuli, regardless of whether the stimulus is low-level (e.g., visual, auditory) or high-level (e.g., emotions; Wilcox et al., 2016). However, this theory contrasts another prominent theory that suggests that migraineurs have a heightened sensitivity to emotionally arousing stimuli, regardless of if the emotional valence is positive or negative (Steppacher et al., 2016). Several studies have also supported the theory of enhanced responses to higher arousal stimuli in migraine, including an electroencephalography study investigating the response to emotional pictures (Steppacher et al., 2016), and studies investigating visual (Sezai et al., 2023) and auditory (Masson et al., 2020) hypersensitivities to low-level arousing stimuli. This enhanced response to emotional stimuli could potentially contribute to the psychiatric comorbidities often observed in migraineurs, such as anxiety and depression (Breslau & Davis, 1993). However, it remains uncertain which of these theories (i.e., a general hypersensitivity to negative stimuli or a heightened sensitivity to emotionally arousing stimuli) may underlie this phenomenon and its effect on multisensory integration in migraineurs.

While the aforementioned research has provided crucial insights into the relationship between migraine, audiovisual multisensory integration, and emotional valence/arousal, the precise mechanisms underlying these interactions remain largely unexplored. Naturalistic stimuli, such as audiovisual films, podcasts, and virtual reality, show promise for exploring how heightened sensitivity to emotional stimuli in migraine may impact multisensory integration.

Naturalistic stimuli are rich, multimodal, and dynamic, and better represent our daily lived experience (Sonkusare et al., 2019). Consequently, naturalistic stimuli can evoke brain responses that are highly reproducible within and across subjects (Hasson et al., 2004). Importantly, naturalistic paradigms engage a broader set of brain regions and more diverse modes of network interactions than task or resting-state paradigms (Zhang et al., 2021). Of particular note, Finn et al. (2018) used naturalistic stimuli to characterize differences in brain synchrony between participants who varied based on personality traits, in this case low and high trait paranoia. Their results showed characteristic patterns of neural synchrony unique to the high paranoia group, suggesting that underlying traits can influence the processing of naturalistic stimuli in the brain. Further, naturalistic stimuli allow for the modulation of emotional valence and arousal, providing a means to explore differences in neural responses related to the higher-level emotional content of the stimulus. Therefore, using naturalistic stimuli will offer insight into neurological processing that is more akin to real-life experience, allowing for a more comprehensive understanding of the interplay between the emotional content of external stimuli, migraine, and multisensory integration.

One common approach for analyzing neural responses to naturalistic stimuli is intersubject correlation analysis (ISC). ISC examines the correlation of hemodynamic responses to an audiovisual stimulus over time across multiple subjects. This approach helps identify neural synchrony, which is the shared neural activity among subjects (Hasson et al., 2009). ISC is particularly effective for naturalistic paradigms as it detects shared neural responses with minimal assumptions, as opposed to GLM-based approaches (Finn et al., 2020). ISC isolates the impact of the stimulus, making it easier to link observed neural activity directly to the stimulus. Therefore, ISC is a highly suitable technique for examining how migraine impacts the synchronous neurological responses to complex audiovisual stimuli.

The current study seeks to investigate differences in neural synchrony between migraineurs and controls in response to complex audiovisual stimuli. Additionally, it aims to explore how emotional arousal and valence influence these differences. In doing so, we will evaluate two competing theories that propose migraineurs may be predominantly sensitive either to emotional arousal or emotional valence. To do so, we acquired fMRI data from 22 individuals with migraine and probable migraine and 21 healthy controls during the passive viewing of a negative valence, high arousal film, a positive valence, high arousal film, and a neutral valence, low arousal film. We hypothesized that if migraineurs are hypersensitive to high arousal stimuli, that they will exhibit heightened neural synchrony in regions associated with multisensory integration (right pSTG, bilateral SPL/IPS, left MTG, and left frontal cortices) during both high arousal negative and positive film conditions. However, if migraineurs are hypersensitive to negative valence stimuli, we hypothesize that they will show heightened neural synchrony in these same multisensory integration regions only during the negative valence, high arousal film. This research will determine how migraine impacts neural synchrony in response to complex audiovisual stimuli and examine whether arousal or valence more profoundly influences neurological processing.

## Methods

### Participants and Stimuli

We recruited 46 participants to watch three short films during fMRI. We had to restart at least one of the films for 2 participants who were subsequently removed from the study. One participant dropped out during MRI scanning and was removed from the study, resulting in 43 total participants (28 females/15 males, mean age = 27.15, standard deviation = 8.32). All participants had no history of neurological illnesses, no hearing impairments and normal or corrected-to-normal vision. To assess migraine status, participants completed the Mickleborough migraine questionnaire (Mickleborough et al., 2016). This questionnaire includes questions that align with the ICHD-3 (Headache Classification Committee of the International Headache Society, 2013) criteria for migraine without aura, migraine with aura, probable migraine without aura, probable migraine with aura, and chronic migraine. 9 participants met the ICHD-3 criteria for migraine without aura (8 females/1 male, mean age = 27.7), 4 participants met the ICHD-3 criteria for migraine with aura (4 females/0 males, mean age = 27.75), 9 participants met the ICHD-3 criteria for probable migraine without aura (5 females/4 males, mean age = 30.33). As probable migraine without aura has symptom and epidemiologic profiles that overlap with migraine and is considered a prevalent form of migraine (Patel et al., 2004), we included participants who met the ICHD-3 classification for migraine without aura, migraine with aura, and probable migraine without aura in the migraine group, resulting in 22 participants in the migraine group (17 females/5 males, mean age = 28.77). The migraine and control groups were approximately matched on age (*t*(41) = -1.287, *p* = 0.205) and level of education (*t*(41) = 1.147, *p* = 0.258). In ISC analysis, a sample size of 22 migraineurs is sufficient and can be robust due to the focus on shared neural responses across subjects (Nastase et al., 2019) and with 20 subjects per group, the ISC statistic converges close to a large sample ISC statistic with 130 subjects (Pajula & Tohka, 2016). The migraine group averaged 42.14 (standard deviation = 47.66) headaches a year, with the average headache lasting 16.52 hours (standard deviation = 23.26). 21 participants who did not meet the ICHD-3 criteria served as the control group (11 females/10 males, mean age = 25.52). The control group averaged 18.45 (standard deviation = 26.45) headaches a year, with the average headache lasting 1.69 hours (standard deviation = 1.36). All participants in the control group reported headaches of only mild-moderate pain intensity, and 18 participants in the migraine group reported moderate-severe pain intensity. No participants reported having a headache within 48 hours prior to participation. This study was approved by the University of Alberta Research Ethics Board (Pro00120063).

Participants watched three short films of differing emotional arousal and valence during fMRI. The first short film, titled “Wicken” (Hashmic House Films, 2019; a short horror film), was negative emotional valence and high arousal. The second short film, titled “One-Minute Time Machine” (Avery, 2023; a short comedy film), was positive emotional valence and high arousal. The third short film, titled “How to Create an Effective Training Video” (iSpring, 2020; a short tutorial), was neutral emotional valence and low arousal. These films were counterbalanced across participants to ensure any possible order effects were minimized and we verbally checked in with participants before and after each film to confirm they wanted to continue. Following each film, participants completed the Modified Differential Emotions Scale (Fredrickson, 2001) to assess emotional responses, resulting in a 2-3 minute break from watching movies in between films. Participants were presented with 16 emotional categories and asked to make a verbal response on a scale of 1 to 7 of the intensity they felt that emotion over the time course of the film. The verbal response was recorded as a keypress response from the experimenter before moving to the next item. All films were edited using VEED.IO (Veed.io, 2024) to add subtitles, remove opening credits/title scenes and remove closing credits. One additional scene was removed from the negative valence, high arousal film to avoid potential motion artifacts because it contained a jump scare.

### Data Acquisition

Neuroimaging data was acquired on a 3-Tesla Siemens Skyra Magnetom scanner at the University 3T MRI Centre on the University of Lethbridge campus. After an initial localizer scan, a high-resolution three-dimensional anatomical image was collected using a T1-weighted magnetization-prepared rapid gradient echo sequence (192 continuous sagittal slices, slice thickness = 1.0mm, matrix size = 256 x 256, field of view = 256mm, voxel size = 1.0mm x 1.0mm x 1.0mm, TR = 2300ms, TE = 2.26ms, flip angle = 8 degrees). Functional images were acquired using a multiband T2*-sensitive gradient-recalled single-shot echo-planar imaging (EPI) sequence (TR = 1500ms, TE = 34.0ms, voxel size = 2.4mm isotropic, flip angle = 90 degrees, matrix size = 85 x 85, field of view = 204mm, multiband factor = 4).

We acquired three different functional scans, one throughout the duration of each of the films watched by participants (i.e., negative valence/high arousal, positive valence/high arousal, neutral valence/low arousal). Stimuli were presented on a 40” MRI compatible LCD screen and the audio portion of the stimulus was delivered through in-ear MRI compatible headphones. A mirror was fixed to the head coil to allow participants to have a direct view of the screen. The position of the LCD screen was marked on the ground with tape to ensure that the positioning of the screen remained consistent across participants. Prior to the functional scans, we tested the alignment of the mirror and LCD screen to ensure that participants had a direct view of the entire screen.

### Data Preprocessing

Initial preprocessing was performed using fMRIPrep 21.0.1 (Esteban et al., 2019). Full preprocessing details can be found in Supplementary Materials A. Briefly, the T1-weighted images were corrected for intensity non-uniformity, skull-stripped, and spatially normalized to MNI152NLin2009cAsym space using advanced normalization tools (ANTs; Avants et al., 2008). For the functional data, we used fMRIPrep 21.0.11 (Esteban et al., 2019) to apply slice-timing correction, head-motion correction with six degrees of freedom. The BOLD data were co-registered to T1-weighted images, and nuisance regressors (e.g., FD, DVARS, CompCor) were computed. Frames exceeding 0.5mm FD or 1.5 standardized DVARS were flagged as motion outliers. Finally, the BOLD time-series were spatially normalized to standard space using ANTs, with final outputs including both volumetric and surface-based resampling. Additional preprocessing was performed using python code that included spatial smoothing with a 6mm FWHM Gaussian kernel, detection and removal of spikes, denoising through regression using motion parameters, cerebrospinal fluid signal, and polynomial trends.

### Behavioral Questionnaires

After passively viewing each film during the fMRI session, participants completed the Modified Differential Emotions Scale (Fredrickson, 2001) to quantify their emotional experience. This 16-question scale comprehensively measures positive, negative, and neutral emotions, as well as subjective arousal, providing a detailed assessment of participants emotional responses to each film. To ensure that each film elicited the intended arousal level, we compared responses to the ‘interested, concentrated, alert’ item between all films using paired-samples *t*-tests in SPSS (version 27; IBM Corp., 2020).

To ensure that the negative valence film elicited the intended emotional response, we used paired-samples *t*-tests in SPSS (version 27; IBM Corp., 2020) to test differences within participants on the average of their answers to the Modified Differential Emotions Scale items ‘fearful, scared, afraid’ and ‘anxious, tense, nervous’ between all films. To ensure that the positive valence film elicited the intended emotional response, we used paired-samples *t*-tests in SPSS (version 27; IBM Corp., 2020) to test differences within participants on the average of their answers to the Modified Differential Emotions Scale items ‘joyful, happy, amused’ and ‘warmhearted, gleeful, elated’ between all films. We also assessed whether migraineurs and controls differed in their emotional responses to the films. To do so, we conducted independent samples *t*-tests comparing migraine and control groups on arousal, negative valence and positive valence ratings for each film using the Modified Differential Emotions Scale items previously mentioned.

Following each MRI scanning session, participants also completed the Beck Depression Inventory-II (BDI; Beck et al., 2011), the Perceived Stress Scale (PSS; Cohen et al., 2014), and the PANAS-GEN questionnaire (Watson et al., 1988). To assess group differences in depression, perceived stress, and positive and negative affect, we conducted independent samples *t*-tests on scores using SPSS (version 27; IBM Corp., 2020).

### Intersubject Correlation Analyses

ISC was computed for all unique pairs of participants using 3dTcorrelate by AFNI (Cox & Hyde, 1997) which produced 903 (*n*\*(*n*-1)/2, where n = 43) unique ISC maps overall. Of these, 231 maps originated from the migraine group (*n*\*(*n*-1)/2, where n = 22), 210 originated from the control group (*n*\*(*n*-1)/2, where n = 21), and 462 ISC maps were obtained from between-group pairings. ISC is a model-free approach used to analyze complex fMRI data acquired in naturalistic, audiovisual stimulus environments (Kauppi, 2010). It allows us to measure shared content across experimental conditions by filtering out subject-specific signals and revealing voxels with a consistent, stimulus-evoked response time series across subjects (Nastase et al., 2019a). It does this by calculating pairwise correlation coefficients between all pairs of participants for each voxel throughout the brain. Ultimately, it provides us with a map of neural synchrony, indicating regions with synchronous ISC across participants throughout the entire time course of the film.

### Linear Mixed Effects Modelling

After running ISC for all pairs of participants, we used the Linear Mixed Effects Modelling (LME) implemented via the *3dISC* module in AFNI (Cox & Hyde, 1997; described by Chen et al., 2020) to identify significant ISC associated with our contrasts of interest. LME is a parametric method based on the general linear model (GLM) and is used to model the relationship between the fMRI time series data and the experimental conditions at each voxel, accounting for the complex covariance structure of ISC data. As LME accounts for variability in the data due to random effects, it allows for more precise estimation of fixed effects (Koerner & Zhang, 2017). The random effects specified in the LME model indicate random intercepts for each subject. These random effects account for individual differences in how participants respond to the same movie stimulus. LME was run separately for each condition (i.e., the audiovisual films) resulting in the following contrasts: comparison of ISC between migraineurs and controls during the negative valence, high arousal film; comparison of ISC between migraineurs and controls during the positive valence, high arousal film; and comparison of ISC between migraineurs and controls during the neutral valence, low arousal film. To address the imbalance in sex distribution between the migraine and control groups, we included sex as a quantitative explanatory variable in our LME model. We used sex as a main effect to account for its potential influence on the results. Random effects for subjects were also included to account for individual variability, allowing us to isolate the effects of interest more accurately. Significant ISCs were defined using a voxelwise false discovery rate (FDR) of *q* < 0.05. Significant results were transformed into surface space for visualization purposes only.

## Results

### Behavioral Results

After passively viewing each film during the fMRI session, participants completed the Modified Differential Emotions Scale (Fredrickson, 2001) to quantify their emotional experience. To ensure that each film elicited the intended arousal level, we used paired-samples *t*-tests in SPSS (version 27; IBM Corp., 2020) to test differences on the Modified Differential Emotions Scale item ‘interested, concentrated, alert’ between all films. There were no significant differences found between negative (mean = 4.907, standard deviation = 1.428) and positive (mean = 5.116, standard deviation = 1.418; *t*(42) = 0.802, *p* = 0.427) films. There was a significant difference found between negative and neutral (mean = 3.279, standard deviation = 1.723; *t*(42) = 5.185, *p* < 0.001), and positive and neutral (*t*(42) = 5.468, *p* < 0.001) films, suggesting that the positive and negative high arousal films elicited significantly higher arousal levels than the neutral, low arousal film.

To ensure that the negative valence film elicited the intended negative emotional response, we averaged the responses from the Modified Differential Emotions Scale items ‘fearful, scared, afraid’ and ‘anxious, tense, nervous’ and used paired-samples *t*-tests in SPSS to test for differences between the three films. There was a significant difference found between negative (mean = 4.778, standard deviation = 1.329) and positive (mean = 1.256, standard deviation = 0.581; *t*(42) = 16.795, *p* < 0.001), and negative and neutral (mean = 1.081, standard deviation = 1.329; *t*(42) = 17.453, *p* < 0.001) films. There was no significant difference found between positive and neutral films (*t*(42) = 1.801, *p* = 0.079). To ensure that the positive valence film elicited the intended emotional response, we used paired-samples *t*-tests in SPSS to test differences within participants on the average of their answers to the Modified Differential Emotions Scale items ‘joyful, happy, amused’ and ‘warmhearted, gleeful, elated’ between all films. There was a significant difference found between positive (mean = 4.884, standard deviation = 1.262) and negative (mean = 1.361, standard deviation = 0.601; *t*(42) = 15.528, *p* < 0.001), positive and neutral (mean = 1.942, standard deviation = 1.221; *t*(42) = 10.658, *p* < 0.001) and neutral and negative (*t*(42) = -3.019, *p* = 0.004), suggesting that the positive film elicited significantly higher positive emotions.

Next, we assessed whether migraineurs and controls differed in their emotional responses to the films. To do so, we conducted independent samples *t*-tests comparing migraine and control groups on the arousal and valence Modified Differential Emotions Scale response previously mentioned. Migraineurs did not differ from controls on arousal during the negative film (*t*(41) = - 0.737, *p* = 0.466), the positive film (*t*(41) = -0.221, *p* = 0.826) or the neutral film (*t*(41) = 0.375, *p* = 0.710). See Figure 1 for the means and standard deviations of the arousal modified differential scores across groups.

**Figure 1.**
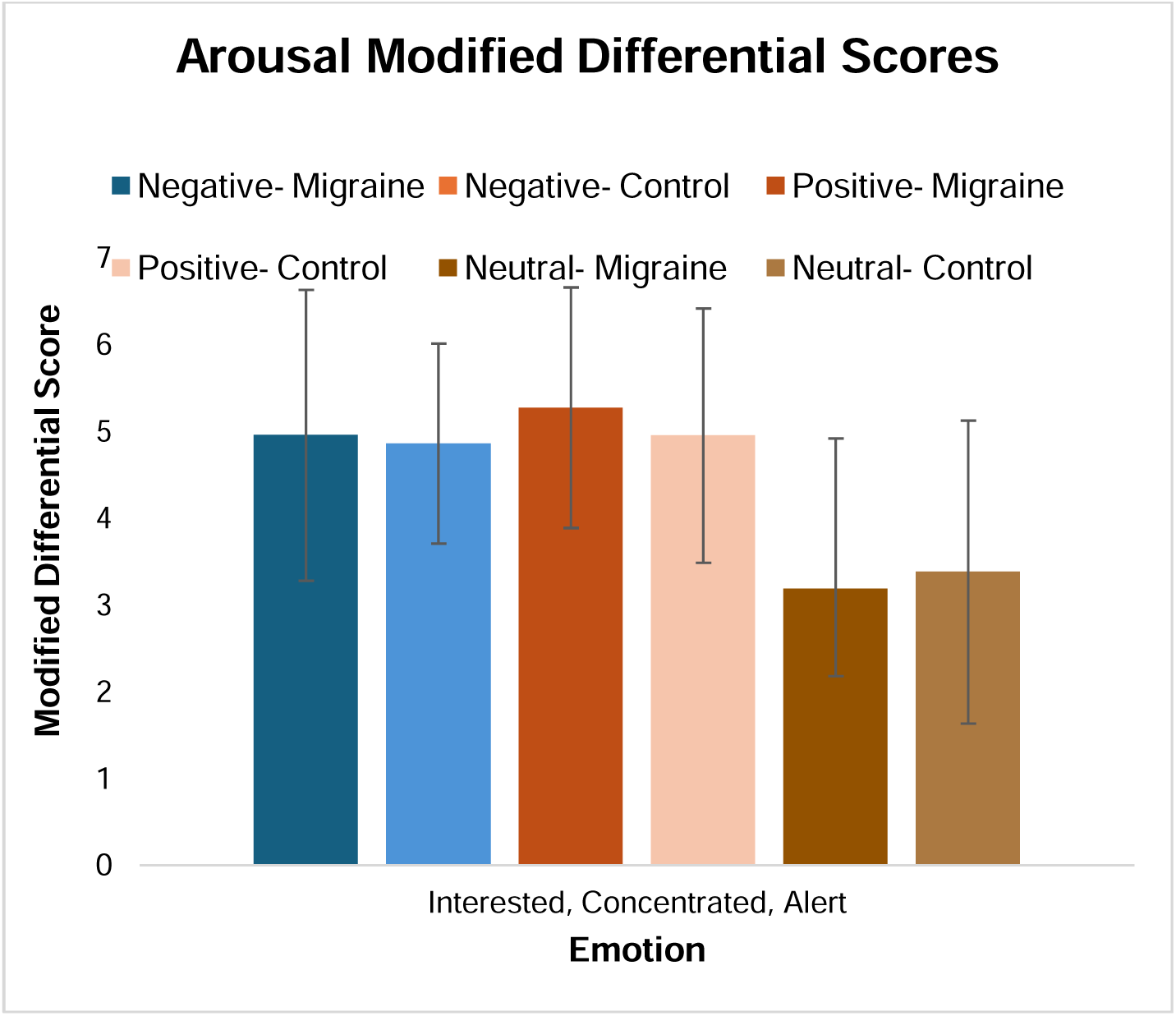
Arousal Modified Differential Scores. *Note.* Means and standard errors for the perceived arousal of each movie type in migraine and control groups. Negative- Migraine: migraine group’s response to the negative valence, high arousal film. Negative- Control: control group’s response to the negative valence, high arousal film. Positive- Migraine: migraine group’s response to the positive valence, high arousal film. Positive- Control: control group’s response to the positive valence, high arousal film. Neutral- Migraine: migraine group’s response to the neutral valence, low arousal film. Neutral- Control: control group’s response to the neutral valence, low arousal film.

For negative valence ratings, migraineurs differed from controls during the negative valence film (*t*(41) = -2.117, *p* = 0.040), but did not differ during the positive (*t*(41) = -0.193, *p* = 0.848) or neutral films (*t*(41) = 1.214, *p* = 0.232). For positive valence ratings, migraineurs did not differ from controls during the negative (*t*(41) = -0.035, *p* = 0.972), positive (*t*(41) = -0.980, *p* = 0.33), or neutral films (*t*(41) = -0.943, *p* = 0.351). Thus, migraineurs and controls did not differ in their emotional responses in terms of arousal or positive valence for any of the films, but they did differ in their negative emotional response during the negative film. Refer to Figure 2 for the means and standard deviations of the valence modified differential scores across groups.

**Figure 2.**
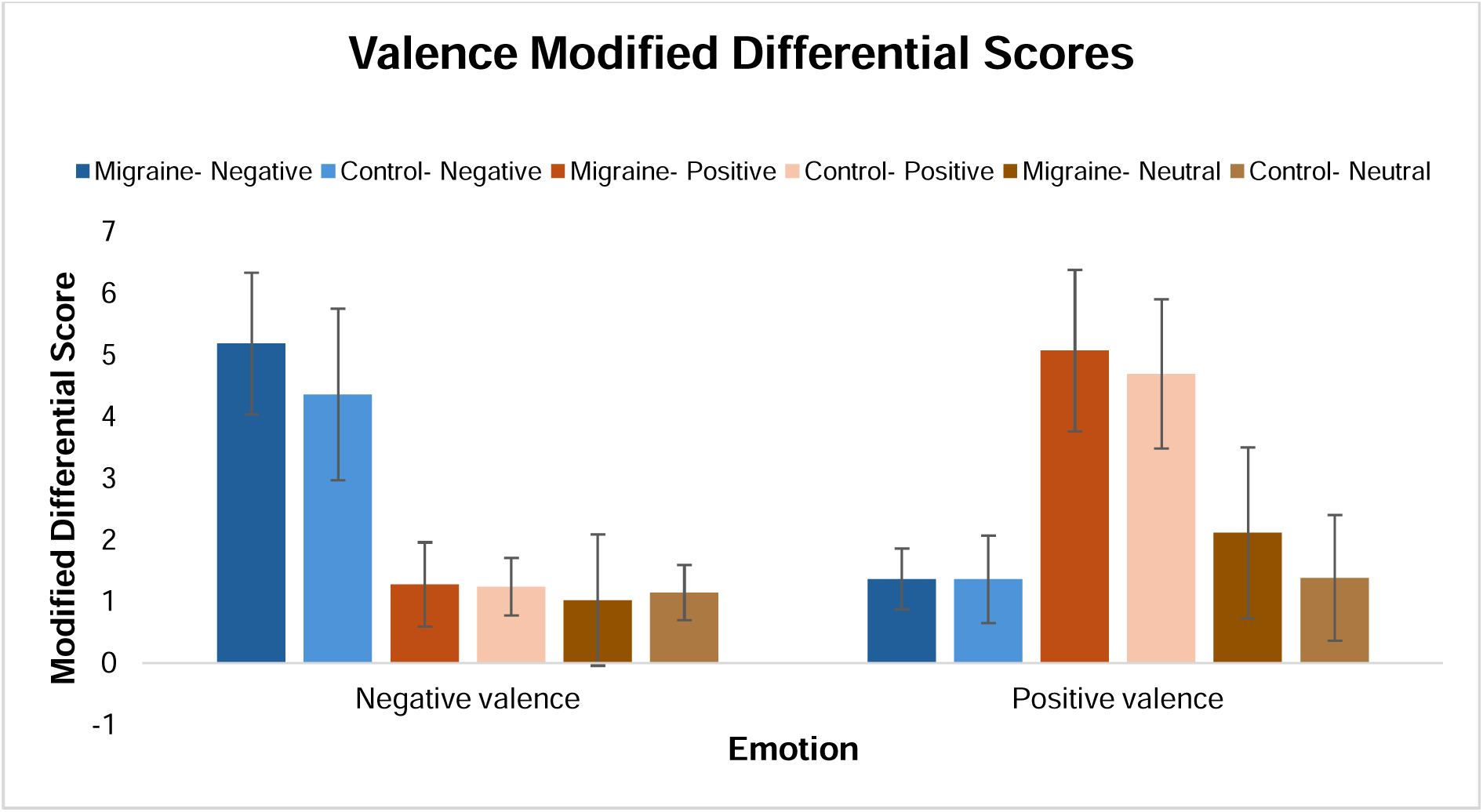
Valence Modified Differential Scores. *Note.* Means and standard errors for the perceived negative and positive valence of each movie type in migraine and control groups. Negative- Migraine: migraine group’s response to the negative valence, high arousal film. Negative- Control: control group’s response to the negative valence, high arousal film. Positive- Migraine: migraine group’s response to the positive valence, high arousal film. Positive- Control: control group’s response to the positive valence, high arousal film. Neutral- Migraine: migraine group’s response to the neutral valence, low arousal film. Neutral- Control: control group’s response to the neutral valence, low arousal film.

Following each MRI scanning session, participants completed the Beck Depression Inventory-II (BDI; Beck et al., 2011), the Perceived Stress Scale (PSS; Cohen et al., 2014), and the PANAS-GEN questionnaire (Watson et al., 1988). To assess group differences in depression, perceived stress, and positive and negative affect, we conducted independent samples t-tests on participants’ scores using SPSS (version 27; IBM Corp., 2020). There were no significant differences in BDI scores (*t*(41) = -1.899, *p* = 0.065), PSS scores (*t*(41) = -1.798, *p* = 0.079), or PANAS-GEN scores (*t*(41) = -0.643, *p* = 0.521).

### Results from LME

Results from the LME examining group differences in processing the negative valence, high arousal film are shown in Figure 3 and Tables S1 and S2 (Supplementary Materials B).

**Figure 3.**
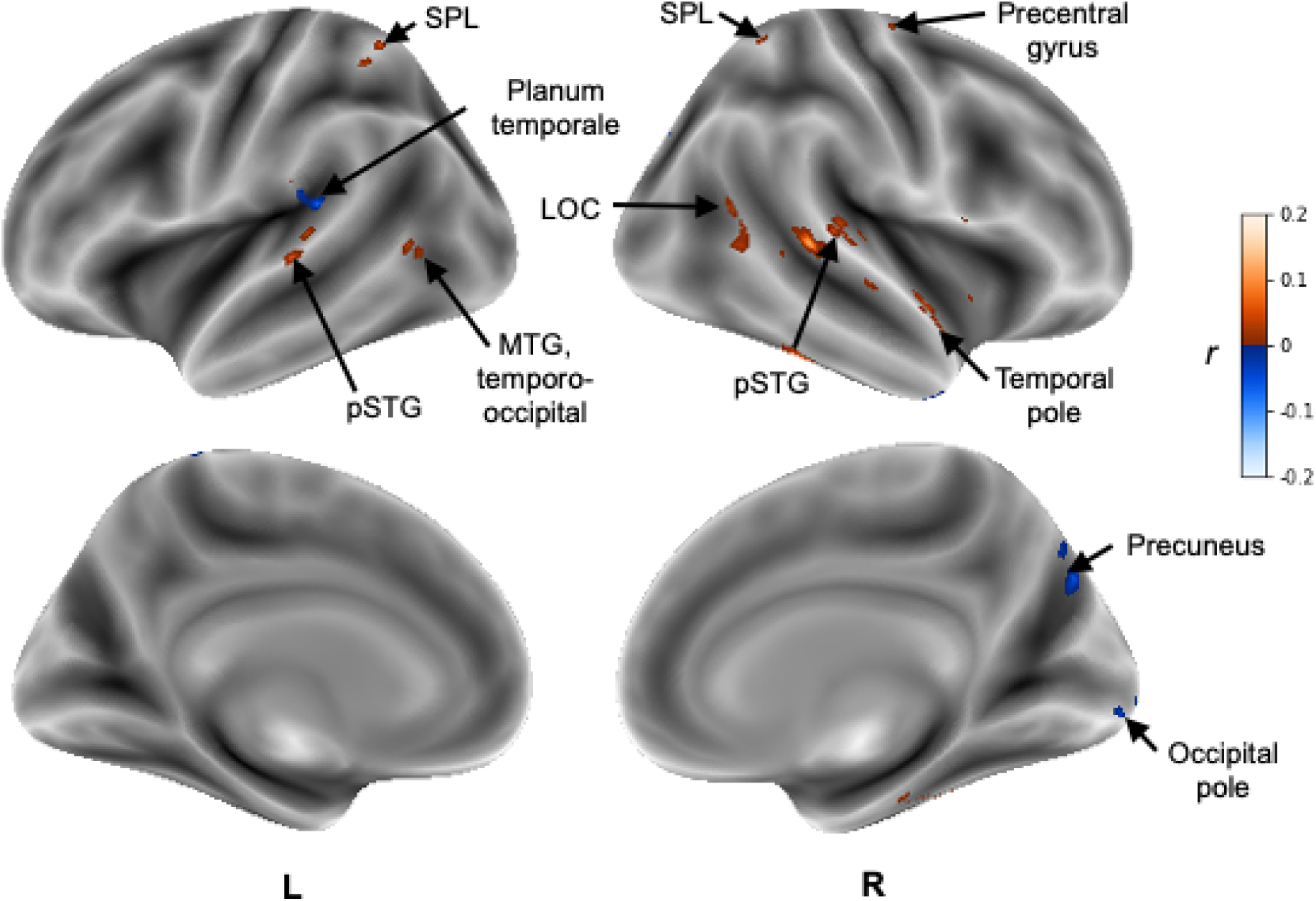
Voxels showing significant ISC within participants with migraine and probable migraine compared to healthy controls across the time course of the negative valence, high arousal audiovisual stimulus. Warm colors refer to regions that are more synchronous in the migraine than the control group. Cool colors refer to regions that are more synchronous in the control than the migraine group. Results are displayed as a voxelwise false discovery (FDR) threshold of *q* < 0.05.

Migraineurs showed significantly greater ISCs than healthy controls in the bilateral pSTG, bilateral SPL, and left MFG. Healthy controls showed significantly greater ISCs than migraineurs in the right precuneus and right occipital pole. Results from the LME examining group differences in processing the positive valence, high arousal film and neutral valence, low arousal film found no significant differences.

## Discussion

This study sought to examine differences in ISCs between migraineurs and healthy controls in response to films of differing emotional valence and arousal (i.e., negative valence/high arousal, positive valence/high arousal, neutral valence/low arousal). To do so, we used ISC analysis and LME modelling to determine how neural synchrony varies between the migraine and control groups across the three film conditions that varied based on emotional valence and arousal. Results from this study show that migraineurs exhibit more similar processing in regions associated with audiovisual multisensory integration, including the bilateral pSTG and SPL, while processing negative valence, high arousal audiovisual stimuli, which may reflect an increased sensitivity to negative emotional information in migraine.

Behaviourally, results from the Modified Differential Emotions Scale confirmed that the high arousal films were found to be more arousing than the neutral film and did not differ in arousal from each other. Further, each film used was found to elicit their intended emotional responses in terms of negative, positive, and neutral valence. When comparing the emotional response between groups, there were no significant group differences for arousal during any of the films. There were significant group differences for negative valence emotions, with the migraine group scoring higher during the negative film. These results validate the fMRI findings which suggest that migraineurs may be hypersensitive to negative valence emotional stimuli.

There were no significant group differences for positive valence emotions during any of the films. Additionally, there were no significant group differences for BDI, PSS, or PANAS-GEN scores.

In response to the negative valence, high arousal film, migraineurs showed heightened neural synchrony in the bilateral pSTG and SPL compared to controls, which are key multisensory integration areas. The pSTG is considered a hub of multisensory integration, combining visual and auditory inputs (Ozker et al., 2017), while the SPL is known to integrate auditory and visual speech (Molholm et al., 2006). Additionally, migraineurs showed heightened neural synchrony in the left MTG, a region implicated in multisensory integration and, more specifically, in audiovisual integration during complex non-speech stimuli (Gao et al., 2023), which is consistent with the type of stimuli used in this study. It is well known that migraineurs experience visual and auditory sensitivities, resulting in altered multisensory integration (Yang et al., 2014). As migraineurs may experience an enhanced response to negative valence stimuli (Steppacher et al., 2016), it is possible that multisensory integration is enhanced during negative/aversive emotional states. Importantly, no differences in neural synchrony were observed in the bilateral pSTG or SPL during the neutral valence, low arousal or positive valence, high arousal films. Together, these findings support the theory that migraineurs have a heightened sensitivity to negative valence emotional stimuli (Wilcox et al., 2016) during the interictal period. Therefore, heightened synchrony in the bilateral pSTG, SPL and left MTG during only the negative valence, high arousal film may reflect an increased effort to integrate the increased sensory input from both auditory and visual modalities. This suggests that in migraineurs, amplified sensory processing associated with hyperexcitable cortex during negative emotional states may also increase the demand on multisensory integration processes, however, more research is needed to determine the exact mechanisms underlying this heightened synchrony.

In response to the negative valence, high arousal film, controls had heightened neural synchrony in the left planum temporale and right occipital pole compared to migraineurs. The left planum temporale is involved in early auditory processing (Binder et al., 1996), where it plays a pivotal role in processing auditory and visual linguistic stimuli (Wani, 2024). The right occipital lobe is involved in early visual processing and is considered the primary distributor of visual information that reaches other cortical areas (Tong, 2003). Given the heightened sensitivity to auditory and visual stimuli in migraine, the increased synchrony observed in these regions in the control group during negative valence audiovisual processing may reflect more stable and uniform early-stage processing, however more research is needed to fully understand the implications of heightened neural synchrony in these regions.

This study sought to examine differences in how migraineurs and controls process naturalistic audiovisual stimuli of differing emotional arousal and valence, however, there are several limitations to be discussed. The films used in this study were chosen to elicit specific valence and arousal responses but were not matched in terms of visual and auditory characteristics. For example, the negative valence, high arousal film had dark lighting, a suspenseful musical score, and less dialogue compared to the positive valence film. These differences may have influenced neural synchrony independently of emotional content, making it difficult to isolate the effects of valence and arousal. Therefore, we did not directly compare across films, as we cannot control for these differences between stimuli. Cross-film comparisons would be challenging to interpret accurately because differences in neural synchrony may result from varying visual and auditory elements rather than emotional valence or arousal alone. Future research should use stimuli where the visual and auditory components are more closely matched, such as using films with similar lighting, color grading, sound design, and dialogue length, which would ensure that only emotional valence differs between conditions and enable direct comparisons across films.

Additionally, based on our sample size, we were unable to examine differences in neural synchrony associated with different subtypes of migraine (e.g., migraine with and without aura). Future research should extend this paradigm to larger samples of each subtype to understand how different migraine pathologies impact the processing of naturalistic stimuli. We also did not assess anxiety levels. Future studies should collect data on trait anxiety to account for its potential influence on ISC results. We also did not collect information about which stage in the migraine cycle each participant was in at the time of the study or medication use, which may be relevant as different phases are associated with sensory processing changes, particularly in the preictal phase (Mykland et al., 2019). Future studies should account for these factors.

In conclusion, this study demonstrates that migraineurs exhibit greater neutral synchrony when processing negative emotional stimuli, particularly in the bilateral pSTG and SPL, suggesting migraineurs have altered multisensory integration in response to negative emotional content. In other words, the results suggest that negative emotional valence, but not high emotional arousal (regardless of valence), alter processing in migraine as migraineurs exhibited heightened neural synchrony in regions linked to audiovisual multisensory integration when responding to negative valence audiovisual stimuli. Overall, this study highlights how migraine uniquely shapes the brain’s response to negative emotional valence, revealing distinct but interconnected differences in neural processing. These findings underscore the broader implications of migraine-related sensory hypersensitivity, particularly in the context of emotionally salient stimuli, and provide valuable insight into how heightened valence sensitivity may drive differences in multisensory integration and visual information processing.

## Data Availability

All data produced in the present study are available upon reasonable request to the authors

## List of abbreviations

pSTG: posterior superior temporal gyrus
SPL: superior parietal lobule
IPS: intraparietal sulcus
MTG: middle temporal gyrus
ISC: intersubject correlation analysis
fMRI: functional magnetic resonance imaging
EPI: echo planar imaging
T1w: T1 weighted
ANTs: advanced normalization tools
LME: linear mixed effects
GLM: general linear model
LOC: lateral occipital cortex
IFG: inferior frontal gyrus
FDR: false discovery rate
SMA: supplementary motor area

## Declarations

### Ethics approval and consent to participate

Ethics approval was obtained from the University of Alberta Research Ethics Board (Pro00120063). Written informed consent was obtained from all participants prior to their participation in the study.

### Consent for publication

Not applicable.

### Availability of data and materials

The datasets used and/or analysed during the current study are available from the corresponding author on reasonable request.

### Competing interests

The authors declare that they have no competing interests.

### Funding

This work was supported by a NSERC Discovery Grant and NSERC Discovery Launch Supplement to C.E. (RGPIN-2021-03568 and DGECR-2021-00297) and an NSERC PGS-D Scholarship to K.K. (Application No: PGS D – 589897 – 2024). The funding source had no involvement in the study design; in the collection, analysis and interpretation of data; in the writing of the report; and in the decision to submit the article for publication.

### Author contributions

KK wrote and prepared the original draft of the manuscript, conceptualized the study, completed formal analysis, and was involved in data acquisition. JC completed review and editing of the manuscript, was involved in analysis of data and data acquisition. CH was involved in methodology planning and data acquisition. KS was involved in methodology planning and data acquisition. PS was involved in data acquisition. CE was involved in conceptualization, data curation, funding acquisition, project administration, review and editing, resources, and supervision. All authors read and approved the final manuscript.

## Acknowledgements

Not applicable.

## Notes

### Competing Interest Statement

The authors have declared no competing interest.

### Funding Statement

This work was supported by a NSERC Discovery Grant and NSERC Discovery Launch Supplement to C.E. and an NSERC PGS-D Scholarship to K.K.

### Author Declarations

Ethics committee/IRB of University of Alberta gave ethical approval for this work

### Summary of Updates

We applied ICHD-3 criteria to all participants uniformly instead of using self-report as a method of migraine group classification. We then re-ran our analyses using our newly defined migraine groups.

