## Supplementary Materials for "Differential fMRI neural synchrony associated with migraine during naturalistic stimuli with negative emotional valence"

**Supplementary Materials A**

Results included in this manuscript come from preprocessing performed using *fMRIPrep*21.0.1 (Esteban, Markiewicz, et al. (2018); Esteban, Blair, et al. (2018); RRID:SCR_016216), which is based on *Nipype* 1.6.1 (K. Gorgolewski et al. (2011); K. J. Gorgolewski et al. (2018); RRID:SCR_002502).
**Anatomical Preprocessing**

A total of 1 T1-weighted (T1w) images per subject were found within the input BIDS dataset. The T1-weighted (T1w) image was corrected for intensity non-uniformity (INU) with N4BiasFieldCorrection (Tustison et al. 2010), distributed with ANTs 2.3.3 (Avants et al. 2008, RRID:SCR_004757), and used as T1w-reference throughout the workflow. The T1w-reference was then skull-stripped with a *Nipype* implementation of the antsBrainExtraction.sh workflow (from ANTs), using OASIS30ANTs as target template. Brain tissue segmentation of cerebrospinal fluid (CSF), white-matter (WM) and gray-matter (GM) was performed on the brain-extracted T1w using fast (FSL 6.0.5.1:57b01774, RRID:SCR_002823, Zhang, Brady, and Smith 2001). Volume-based spatial normalization to one standard space (MNI152NLin2009cAsym) was performed through nonlinear registration with antsRegistration (ANTs 2.3.3), using brain-extracted versions of both T1w reference and the T1w template. The following template was selected for spatial normalization: *ICBM 152 Nonlinear Asymmetrical template version 2009c*[Fonov et al. (2009), RRID:SCR_008796; TemplateFlow ID: MNI152NLin2009cAsym].

**Functional Data Preprocessing**

For each of the 3 EPI image series found per subject (across all tasks and sessions), the following preprocessing was performed. First, a reference volume and its skull-stripped version were generated using a custom methodology of *fMRIPrep*. Head-motion parameters with respect to the BOLD reference (transformation matrices, and six corresponding rotation and translation parameters) are estimated before any spatiotemporal filtering using mcflirt (FSL 6.0.5.1:57b01774, Jenkinson et al. 2002). BOLD runs were slice-time corrected to 0.69s (0.5 of slice acquisition range 0s-1.38s) using 3dTshift from AFNI (Cox and Hyde 1997, RRID:SCR_005927). The BOLD time-series (including slice-timing correction when applied) were resampled onto their original, native space by applying the transforms to correct for head-motion. These resampled BOLD time-series will be referred to as *preprocessed BOLD in original space*, or just *preprocessed BOLD*. The BOLD reference was then co-registered to the T1w reference using mri_coreg (FreeSurfer), followed by flirt (FSL 6.0.5.1:57b01774, Jenkinson and Smith 2001) with the boundary-based registration (Greve and Fischl 2009) cost-function. Co-registration was configured with six degrees of freedom. Several confounding time-series were calculated based on the *preprocessed BOLD*: framewise displacement (FD), DVARS and three region-wise global signals. FD was computed using two formulations following Power (absolute sum of relative motions, Power et al. (2014)) and Jenkinson (relative root mean square displacement between affines, Jenkinson et al. (2002)). FD and DVARS are calculated for each functional run, both using their implementations in *Nipype* (following the definitions by Power et al. 2014). The three global signals are extracted within the CSF, the WM, and the whole-brain masks. Additionally, a set of physiological regressors were extracted to allow for component-based noise correction (*CompCor*, Behzadi et al. 2007). Principal components are estimated after high-pass filtering the *preprocessed BOLD* time-series (using a discrete cosine filter with 128s cut-off) for the two *CompCor* variants: temporal (tCompCor) and anatomical (aCompCor). tCompCor components are then calculated from the top 2% variable voxels within the brain mask. For aCompCor, three probabilistic masks (CSF, WM and combined CSF+WM) are generated in anatomical space. The implementation differs from that of Behzadi et al. in that instead of eroding the masks by 2 pixels on BOLD space, the aCompCor masks are subtracted a mask of pixels that likely contain a volume fraction of GM. This mask is obtained by thresholding the corresponding partial volume map at 0.05, and it ensures components are not extracted from voxels containing a minimal fraction of GM. Finally, these masks are resampled into BOLD space and binarized by thresholding at 0.99 (as in the original implementation). Components are also calculated separately within the WM and CSF masks. For each CompCor decomposition, the *k* components with the largest singular values are retained, such that the retained components’ time series are sufficient to explain 50 percent of variance across the nuisance mask (CSF, WM, combined, or temporal). The remaining components are dropped from consideration. The head-motion estimates calculated in the correction step were also placed within the corresponding confounds file. The confound time series derived from head motion estimates and global signals were expanded with the inclusion of temporal derivatives and quadratic terms for each (Satterthwaite et al. 2013). Frames that exceeded a threshold of 0.5 mm FD or 1.5 standardised DVARS were annotated as motion outliers. The BOLD time-series were resampled into standard space, generating a *preprocessed BOLD run in MNI152NLin2009cAsym space*. First, a reference volume and its skull-stripped version were generated using a custom methodology of *fMRIPrep*. All resamplings can be performed with *a single interpolation step* by composing all the pertinent transformations (i.e. head-motion transform matrices, susceptibility distortion correction when available, and co-registrations to anatomical and output spaces). Gridded (volumetric) resamplings were performed using antsApplyTransforms (ANTs), configured with Lanczos interpolation to minimize the smoothing effects of other kernels (Lanczos 1964). Non-gridded (surface) resamplings were performed using mri_vol2surf (FreeSurfer).

**Supplementary Materials B**

**Table S1**

*Coordinates and cluster sizes for higher ISC in the probable migraine group compared to the control group in response to the negative valence, high arousal film.*

| **Anatomical Location** | **Hemisphere** | **# of Voxels** | **MAX ISC** | **MAX x** | **MAX y** | **MAX z** |
| --- | --- | --- | --- | --- | --- | --- |
| Superior Temporal Gyrus, posterior division | Right | 51 | 0.2 | 58 | -32 | 6 |
| Temporal Occipital Fusiform Cortex | Right | 25 | 0.133 | 44 | -40 | -22 |
| Planum Polare | Right | 11 | 0.117 | 50 | 0 | -6 |
| Lateral Occipital Cortex, inferior division | Right | 9 | 0.166 | 54 | -64 | 10 |
| Superior Parietal Lobule | Left | 5 | 0.125 | -34 | -58 | 66 |
| Superior Parietal Lobule | Right | 4 | 0.119 | 30 | -52 | 70 |
| Superior Temporal Gyrus, posterior division | Left | 4 | 0.168 | -68 | -24 | 4 |
| Superior Temporal Gyrus, posterior division | Right | 3 | 0.129 | 56 | -18 | -2 |
| Superior Temporal Gyrus, posterior division | Right | 3 | 0.117 | 48 | -26 | 4 |
| Superior Parietal Lobule | Left | 3 | 0.12 | -42 | -50 | 58 |
| Precentral Gyrus | Right | 2 | 0.107 | 24 | -12 | 70 |
| Temporal Pole | Right | 2 | 0.0951 | 50 | 10 | -10 |
| Lateral Occipital Cortex, inferior division | Left | 2 | 0.134 | -52 | -64 | 6 |
| Superior Temporal Gyrus, posterior division | Right | 2 | 0.146 | 66 | -28 | 8 |
| Planum Temporale | Left | 2 | 0.143 | -60 | -30 | 10 |
| Temporal Pole | Right | 1 | 0.109 | 58 | 10 | -4 |
| Middle Temporal Gyrus, temporooccipital part | Left | 1 | 0.108 | -46 | -60 | 4 |
| Lateral Occipital Cortex, inferior division | Right | 1 | 0.142 | 48 | -60 | 14 |
| Temporal Fusiform Cortex, posterior division | Right | 1 | 0.0755 | 32 | -26 | -28 |
| Left VIIb | Left | 1 | 0.0774 | -38 | -42 | -50 |

**Table S2**

*Coordinates and cluster sizes for higher ISC in the control group compared to the probable migraine group in response to the negative valence, high arousal film.*

| **Anatomical Location** | **Hemisphere** | **# of Voxels** | **MAX ISC** | **MAX x** | **MAX y** | **MAX z** |
| --- | --- | --- | --- | --- | --- | --- |
| Precuneus | Right | 12 | 0.124 | 20 | -74 | 34 |
| Planum Temporale | Left | 8 | 0.144 | -48 | -38 | 16 |
| Occipital Pole | Right | 4 | 0.134 | 10 | -98 | -4 |
| Postcentral Gyrus | Left | 3 | 0.0974 | -10 | -44 | 78 |
| Temporal Pole | Right | 1 | 0.0803 | 42 | 4 | -42 |
| Lateral Occipital Cortex, superior division | Right | 1 | 0.107 | 16 | -72 | 42 |
